# Comparison of troponin and natriuretic peptides in takotsubo syndrome and acute coronary syndrome: a meta-analysis

**DOI:** 10.1101/2023.11.21.23298868

**Authors:** Liam S Couch, James W Garrard, John A Henry, Rafail A Kotronias, Bashir Alaour, Giovanni Luigi De Maria, Keith M Channon, Adrian P Banning, Alexander R Lyon, Michael S Marber, Thomas E Kaier

**Affiliations:** King’s College London BHF Centre, The Rayne Institute, St Thomas’ Hospital, London, UK; Division of Cardiovascular Medicine, Radcliffe Department of Medicine, John Radcliffe Hospital, University of Oxford, Oxford, UK; Department of Cardiology, Royal Brompton Hospital, London, UK

**Keywords:** Takotsubo syndrome, acute coronary syndrome, biomarker, troponin, natriuretic peptides

## Abstract

**Background:** Takotsubo syndrome (TTS) is an acute heart failure syndrome which resembles acute coronary syndrome (ACS) at presentation. Differentiation requires coronary angiography, but where this does not occur immediately, cardiac biomarkers may provide additional utility. We performed a meta-analysis to compare troponin and natriuretic peptides (NPs) in TTS and ACS to determine if differences in biomarker profile can aid diagnosis.

**Methods:** We searched five literature databases for studies reporting NPs (BNP/NT-pro-BNP) or troponin I/T in TTS and ACS, identifying 28 studies for troponin/NPs (5618 and 1145 patients respectively).

**Results:** Troponin was significantly lower in TTS than ACS (standardized mean difference (SMD) −0.86; 95% CI −1.08-−0.64; p<0.00001), with an absolute difference of 75 times the upper limit of normal (xULN) higher in ACS than TTS. Conversely, NPs were significantly higher in TTS (SMD 0.62; 0.44-0.80; p<0.00001) and 5.8xULN greater absolutely. Area under the curve (AUC) for troponin in ACS *versus* TTS was 0.82 (0.70-0.93), and 0.92 (0.80-1.00) for STEMI vs. TTS. For NPs, AUC was 0.69 (0.48-0.89). Combination of troponin and NPs with logistic regression did not improve AUC. Recursive partitioning and regression tree analysis calculated a troponin threshold ≥26xULN that identified 95% cases as ACS where and specificity for ACS were 85.71% and 53.57% respectively, with 94.32% positive predictive value and 29.40% negative predictive value.

**Conclusions:** Troponin is lower and NPs higher in TTS *versus* ACS. Troponin had greater power than NPs at discriminating TTS and ACS, and with troponin ≥26xULN patients are far more likely to have ACS.

## Introduction

Takotsubo syndrome (TTS) is a stress-induced acute heart failure syndrome[1] which typically presents with cardiac chest pain and dyspnoea. Electrocardiogram (ECG) abnormalities are common, most frequently manifesting as ST-segment elevation, but may also present as ST-segment depression, widespread T-wave inversion and QTc prolongation.[2] Whilst this classically is not localized to a coronary territory, differentiating from acute coronary syndrome (ACS) based on clinical features alone in the emergency department is often not possible, and diagnosis depends upon invasive or non-invasive coronary angiography, where exclusion of culprit coronary artery stenosis or occlusion is required. Subsequent ventriculography shows typical regional wall motion abnormalities, with a pattern of circumferential apical hypokinesia and basal hypercontractility being the most common anatomical variant. Early studies suggested 1–2% of patients with suspected ACS are eventually diagnosed with TTS,[3,4] although with increasing awareness and more widespread access to early coronary angiography, more recent estimates suggest TTS may constitute up to 5–6% of female patients presenting with suspected ST-segment elevation myocardial infarction (STEMI).[5]

Where patients do not undergo coronary angiography immediately, patients must be treated as ACS. Cardiac biomarkers represent a possible tool in understanding the differences between these two conditions which present similarly. TTS patients exhibit increases in creatine kinase-MB, cardiac troponin (cTn) and natriuretic peptides (NPs),[2] although the release profile appears different from ACS. The literature suggests smaller rises in troponin and creatinine kinase, and higher brain NP in TTS compared to ACS.[6] This likely reflects the modest acute myocyte damage in contrast to the extreme ventricular dilatation and wall stress which occurs in TTS. Indeed, BNP or NT-pro-BNP levels may be extremely high, and elevated NP levels have been included in the updated diagnostic criteria for TTS from the European Society of Cardiology.[1] Admission NT-pro-BNP levels are an independent predictor of 30-day major adverse clinical events and long-term mortality.[7] Consequently, the ratio of NP to troponin has been proposed to differentiate TTS from STEMI.[8] Alternative biomarkers for TTS exist, such as microRNAs and those linked with inflammation and vascular dysfunction, and these have recently been reviewed but are not routinely available for clinical practice.[9–11]

There is a paucity of prospective data in TTS and retrospective studies reporting the levels of established cardiac biomarkers in TTS are often small. To enhance clinical understanding of the differences in biomarker release between ACS and TTS, we performed a meta-analysis of all available literature to compare cardiac troponin and NPs (BNP or NT-pro-BNP) profiles in TTS *versus* ACS and demonstrate a troponin ‘threshold’ above which cases are extremely likely to be ACS.

## Methods

### Search strategy and selection criteria

We identified studies of interest through systematic search of existing literature into TTS and cardiac biomarkers. Five literature databases were searched up to 10^th^ July 2023, including Pubmed, Ovid MEDLINE, EMBASE Ovid, Scopus and Cochrane Central Register of Controlled Trials (CENTRAL) without language, publication year or publication status restrictions. Search terms were adjusted as per individual database requirements but consisted of a stem of “(takotsubo cardiomyopathy OR takotsubo syndrome OR stress induced cardiomyopathy) AND (biomarker OR troponin OR BNP) AND (acute coronary syndrome OR STEMI OR NSTEMI)”. Collated papers were imported into Mendeley Desktop (version 1.19.8), and duplicates were automatically identified and manually removed. This list was incorporated into Excel, where further duplicates were removed, and papers methodically screened for inclusion.

Two review authors (LSC or BA) independently assessed whether the titles and abstracts were eligible for further reading. Articles were read to assess eligibility for inclusion. Any disagreements about study inclusion were resolved through discussion with a third review author (TEK or MM).

Studies that compared TTS to ACS (STEMI, NSTEMI or pooled ACS) for troponin I, troponin T, BNP or NT-pro-BNP were identified for this analysis. Studies must have used either Mayo clinic[12] or European Society of Cardiology criteria[1] for the diagnosis of TTS, or these were retrospectively applied. For inclusion, data must have been displayed as mean and standard deviation (SD) or median and lower/upper quartile. Abstracts, case reports and case series with fewer than 10 patients were excluded from the analysis.

### Data extraction and statistical analysis

Where available, included studies had data extracted for patient number, troponin and/or NP, biomarker assay, diagnostic criteria and reference ranges used. In cases where the biomarker assay was not reported, authors were contacted. Where they did not respond, an averaged value per assay for each isoform was formulated using published upper limits of normal (ULN) for troponin I/T[13] or for NT-pro-BNP/BNP for rule-out of heart failure as per the International Federation of Clinical Chemistry.[14] Peak troponin and BNP/NT-pro-BNP values were used for analysis where specified.

Median and lower/upper quartile was converted to an estimated mean ± SD to facilitate inclusion of the data within the meta-analysis using established methodology produced by Wan et al.[15] Data extracted was normalized to the ULN (xULN) to enable standardized comparison of various biomarker isoforms and assays. Where studies reported NSTEMI and STEMI groups separately against a single TTS control group, NSTEMI and STEMI values were pooled into a single ‘ACS’ group. Analysis of study data was performed by an inverse-variance method with random effects using RevMan 5.[16] Despite standardization, data in forest plots are displayed as standardized mean difference (SMD) with 95% confidence interval due to high study heterogeneity. The presence of heterogeneity amongst studies was assessed with Chi-squared or Higgins and Thompsons’ I^2^ index. Risk of bias analysis was performed using the National Heart, Lung and Blood Institute (NHLBI) National Institute of Health (NIH) Quality Assessment of Case-Control Studies tool[17] by JAH. A planned sensitivity analysis and investigation of heterogeneity based on the study design, sex distribution, timing of blood biomarkers, and overall risk of bias was performed. Statistical significance was determined using a Z-test, with p-value displayed. Recursive Partitioning and Regression Tree analysis was performed for systematic identification of cut-off thresholds using the rpart module in RStudio based on established methodology from Breiman et al.[18]

## Results

### Literature selection

During the initial literature search, 1982 studies were identified using the search strategy outlined above. After removal of duplicates, 1381 unique articles were screened, 28 studies ultimately met criteria for inclusion in this meta-analysis (Figure 1). This enabled comparison of total 5618 patients for troponin and 1145 patients for NPs.2,6,8,16-40

**Figure 1.**
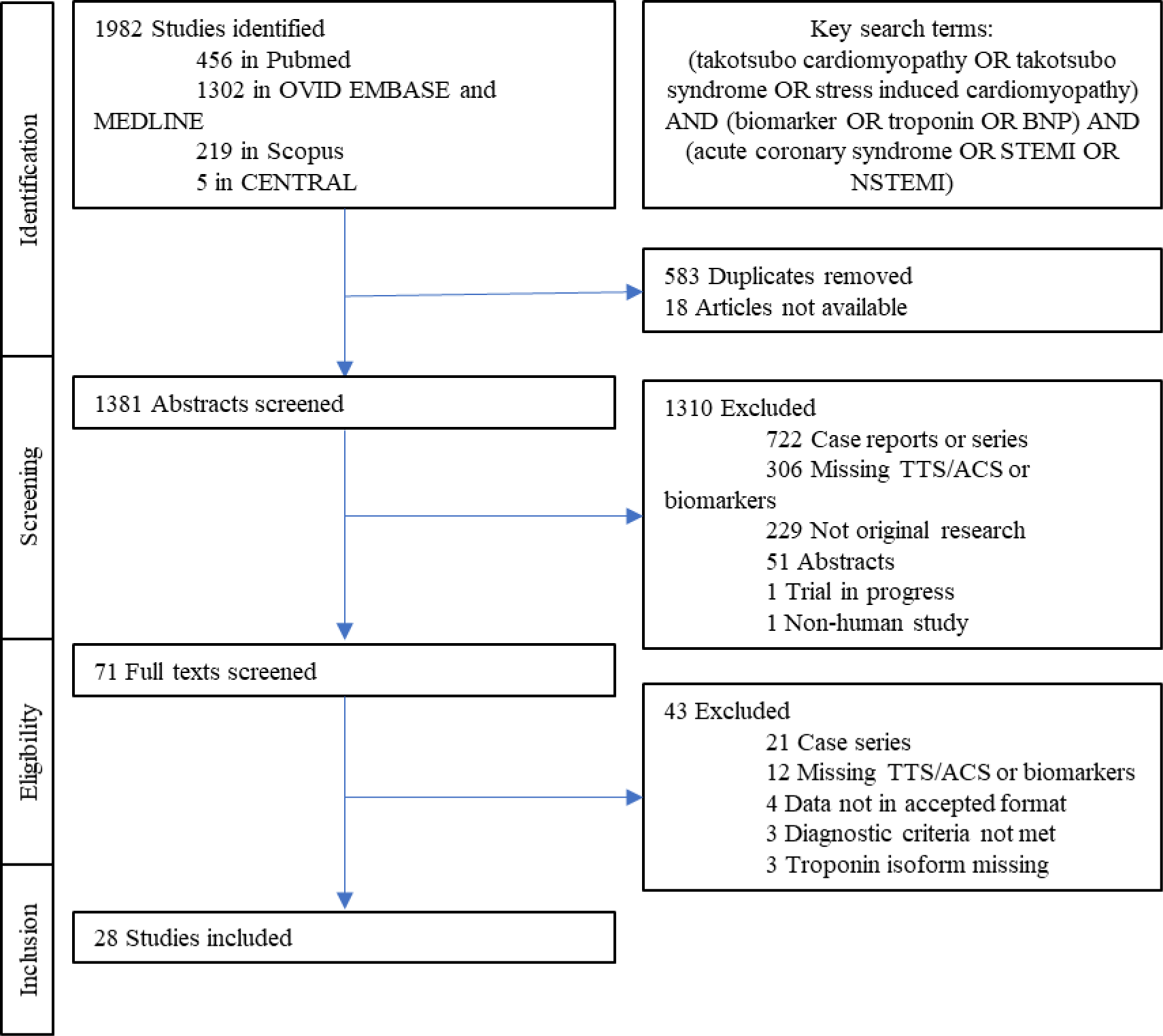
Schematic representation of the workflow for identifying studies Shown is the process involved in performing the meta-analysis, where studies where identified, screened, and then ultimately included or excluded.

### Troponin in TTS versus ACS

Twenty-seven studies compared troponin measurements in TTS and ACS resulting in a total of 1625 patients included for TTS and 3933 for ACS. When normalized to the ULN, troponin was significantly lower in TTS than in ACS (Figure 2A), with a SMD of −0.86xULN (95% CI, −1.08 to −0.64; p<0.00001). Absolutely, troponin was 75.35 times the ULN (95% CI, 57.94 to 92.77) higher in ACS than TTS (p<0.00001; Supplementary Figure 1A).

**Figure 2.**
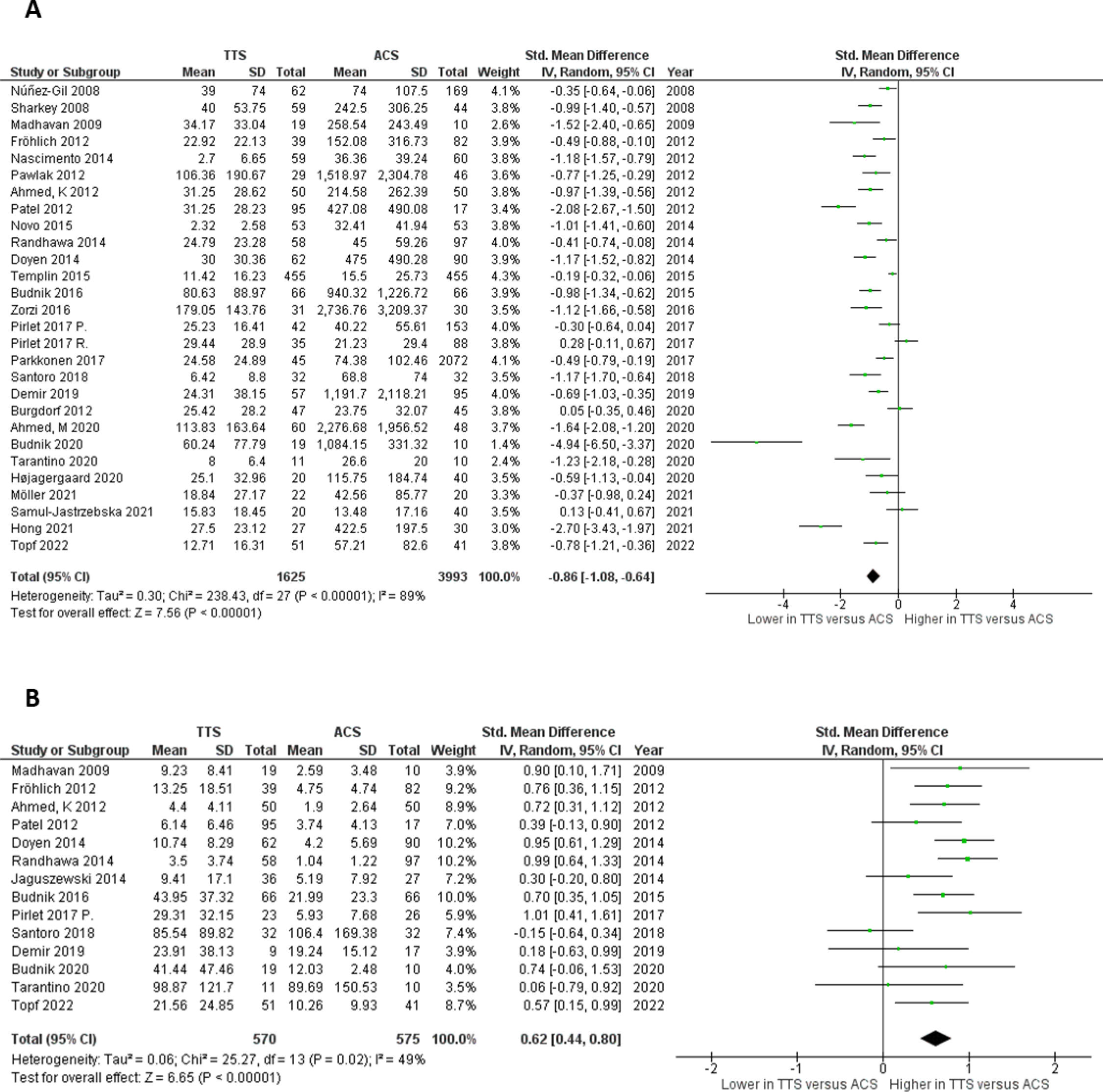
Forest plots for troponin and natriuretic peptide in takotsubo syndrome versus acute coronary syndrome Standardized mean difference shown for included studies in TTS versus ACS for (**A**) troponin and (**B**) NP. Data displayed as mean +/− SD to 2 decimal places. Studies included within this Forest plot: [2,6,8,19–43]. ‘Pirlet 2017 P’ and ‘Pirlet 2017 R’ represent the prospective and retrospective cohorts from this study respectively.

### NPs in TTS versus ACS

NP (BNP and NT-pro-BNP) measurements comparing TTS and ACS was performed from 14 studies, yielding 570 patients with TTS and 575 with ACS. NPs were significantly higher in TTS than ACS (SMD 0.62xULN; 95% CI, 0.44 to 0.80; p<0.00001; Figure 2B) with an absolute difference of 5.88 times the ULN greater in TTS (95% CI, 3.75 to 8.00; p<0.00001; Supplementary Figure 1B).

Subgroup analysis was performed based on NP subtype, BNP or NT-pro-BNP (Supplementary Figure 2). There was a significant difference between the mean differences in BNP and NT-pro-BNP (3.58xULN; 95% CI 2.05 to 5.11; *versus* 13.39xULN; 95% CI 5.97 to 20.81 respectively; p=0.01), and both were significantly higher in TTS than ACS (p<0.00001 and p=0.0004 respectively, Supplementary Figure 2A). However, when standardized to variability, the SMD trended to be greater in BNP than NT-pro-BNP (0.83xULN; 95% CI 0.63 to 1.03; *versus* 0.48xULN; 95% CI 0.19 to 0.76 respectively; P=0.05, Supplementary Figure 2B).

### Subgroup analysis based on type of ACS

A subgroup analysis was performed to assess whether the magnitude of difference in troponin or NP varies based on type of acute MI versus TTS. Troponin measurements (Figure 3A) were significantly lower in TTS than both subgroups (SMD for unspecified ACS/NSTEMI −0.48xULN; 95% CI, −0.69 to −0.27; p<0.00001 and STEMI −1.36xULN; 95% CI, −1.71 to −1.02; p<0.00001). The magnitude of the difference was larger in the STEMI subgroup than for unselected ACS/NSTEMI (p<0.0001).

**Figure 3.**
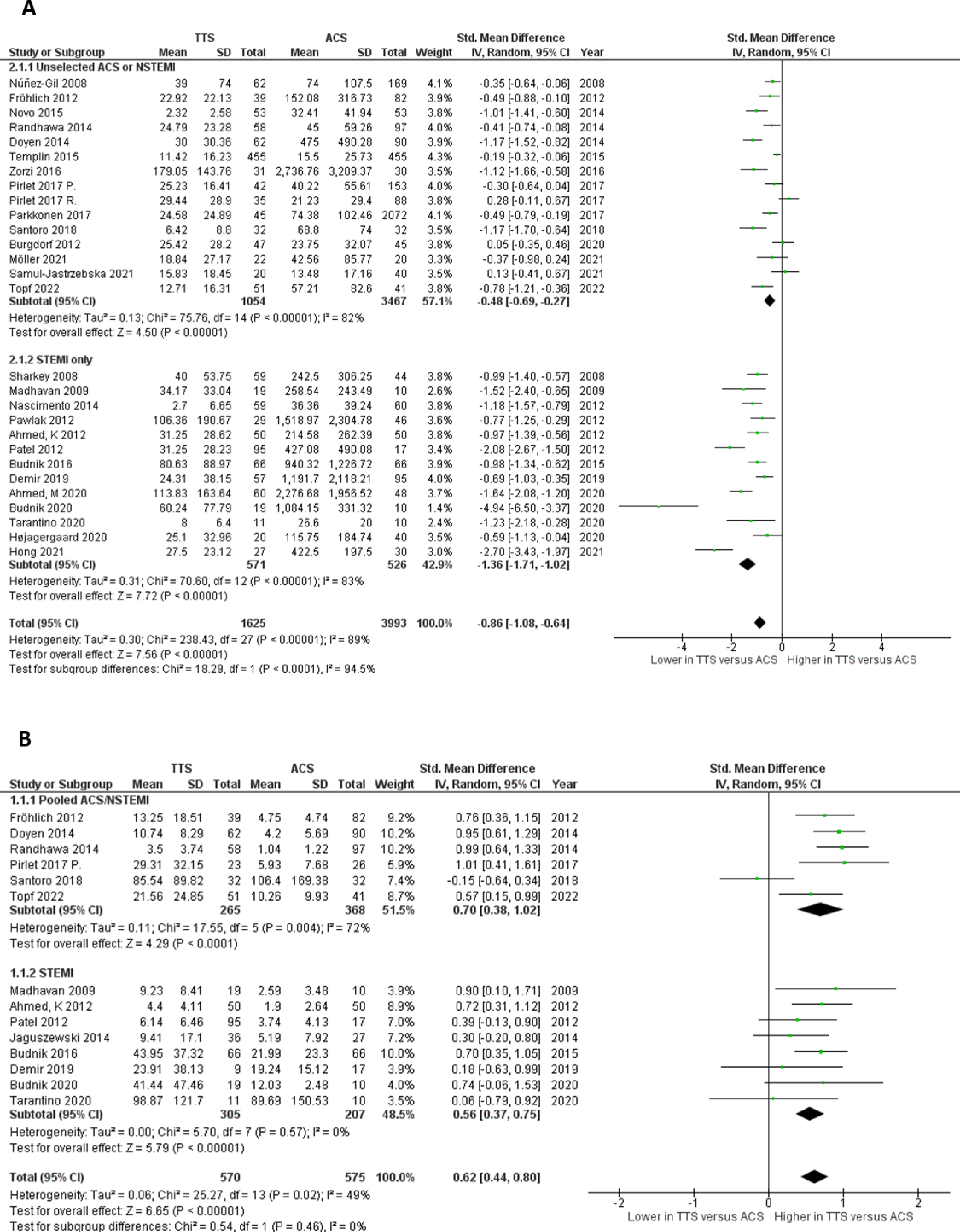
Subgroup analysis comparing the effect of acute myocardial infarction sub-type Standardized mean difference shown stratified by ACS subgroup for (**A**) troponin and (**B**) NP. Data displayed as mean +/− SD to 2 decimal places. Studies included within this Forest plot: [2,6,8,19–43]. ‘Pirlet 2017 P’ and ‘Pirlet 2017 R’ represent the prospective and retrospective cohorts from this study respectively.

NPs (Figure 3B) were significantly higher in TTS than both subgroups (SMD for unspecified ACS/NSTEMI 0.70xULN; 95% CI, 0.38 to 1.02; p <0.0001; and STEMI 0.56xULN; 95% CI, 0.37 to 0.75; p<0.00001). However, there was no difference in the SMD between these two subgroups (p=0.46).

### Receiver operating characteristic curves for troponin and NP for the differentiation of TTS from ACS

Receiver operating characteristic (ROC) curves were constructed based on included studies (Supplementary Figure 3). Since troponin is a sensitive biomarker for myocyte injury and the priority would be to not miss any cases of ACS, TTS was selected for use as the control. Based on included studies, area under the curve (AUC) for troponin in ACS *versus* TTS (Supplementary Figure 3A) was 0.82 (95% CI, 0.70 to 0.93), which enabled good discrimination. Sensitivity was 100% at a troponin value of 13.09 times the ULN, although specificity was 21.43%. A troponin value of 40.11 times the ULN provided sensitivity of 75.00% and specificity of 82.14%. Interestingly, performing ROC analysis on the STEMI subgroup improved AUC to 0.92 (95% CI, 0.80 to 1.00, Supplementary Figure 3B).

ROC analysis revealed a lower AUC (AUC 0.69; 95% CI, 0.48 to 0.89, Supplementary Figure 3C) in the 14 studies including NP measurement. A NP of 3.04 times the ULN provided specificity of 100% and sensitivity of 21.43%, whereas for a NP of 6.03 times the ULN sensitivity was 85.71% and specificity was 57.14%.

The two cardiac biomarkers were combined using a logistic regression model to investigate whether the biomarkers in combination increased the power of discrimination. Here, we only included the 13 studies which recorded both troponin and NPs to enable a paired comparison. In this sub-analysis, the AUC for troponin was 0.92 (95% CI, 0.82 to 1.00) and 0.69 (95% CI, 0.47 to 0.90) for NP (Supplementary Figure 3D and E respectively). After combination, AUC did not improve over troponin alone (AUC = 0.91; 95% CI, 0.79 to 1.00; Supplementary Figure 3F).

### Decision tree analysis to identify ‘optimal’ biomarker threshold

A Recursive Partitioning and Regression Tree analysis was performed to systematically identify biomarker ‘threshold’ values to differentiate TTS from ACS based on the included studies.[18] Given TTS is potentially under diagnosed, we made the conservative assumption that prior to definitive diagnosis, 90% of cases represent MI and 10% account for TTS. Offering both NP and troponin to the decision tree with all available data and favouring ACS as the diagnosis, a value for troponin of 25.92 times the ULN was produced, which when exceeded, defined 95.12% cases as ACS. At this cut-off value, sensitivity and specificity are 85.71% and 53.57% respectively (Supplementary 3A), with a positive predictive value (PPV) of 94.32% and negative predictive value (NPV) of 29.40%.

### Risk of bias, heterogeneity and sensitivity analyses

The risk of bias analysis of included studies was performed using the NHLBI NIH Quality Assessment of Case-Control Studies tool (Supplementary Table 1).[17] Overall, 13 studies were found to be of good quality, 12 studies of fair quality and 2 studies of poor quality.

We conducted a sensitivity analysis of the study designs, sex distribution, risk of bias and timing of blood biomarkers (Supplementary Table 2&3). Excluding poor quality studies, retrospective study design and in studies where timing of biomarker sampling was not given in the study made no significant difference to the point estimate of either the SMD or mean difference for troponin or NP. The sensitivity analysis demonstrated that in studies of only female patients, the SMD of troponin (−1.49; 95% CI −1.99 to −0.99) was lower compared with studies not with only female participants (−0.66; 95% CI −0.88 to −0.45) and lower than the overall cohort. This observation was also seen in the mean difference, with a greater mean difference observed in the female only studies (−1087.48; 95% CI −1548.48 to −626.48). Excluding studies with unequal sex distribution did not affect the SMD or mean difference of either troponin, or NP.

There was a high degree of heterogeneity in the overall analysis, with I^2^ values for SMD of 89% for troponin and 49% for NP (Supplementary Table 2). There was no change in the heterogeneity of the included cohorts based on risk of bias, study design or timing of biomarker. For NP, less heterogeneity was seen in the 4 studies of female only studies (I^2^ = 0%; Supplementary Table 2&3), although there was no meaningful difference in either the SMD or mean difference as described above. In the former analysis of NP subtype (Supplementary Figure 2), heterogeneity was reduced when SMD for BNP and NT-pro-BNP were analyzed separately, with I^2^ values of 9% and 52% respectively.

## Discussion

TTS remains a diagnostic challenge and cardiac biomarkers represent a tool that could aid clinicians when coronary angiography is not performed (or not accessible) acutely. Here we perform a meta-analysis of all available literature in TTS to demonstrate that: (1) troponin is significantly higher in ACS than TTS; (2) NPs are significantly higher in TTS than ACS; (3) on sub-group analysis, expectedly the mean difference in troponin is greater for STEMI than unspecified ACS/NSTEMI *versus* TTS; (4) troponin alone and not in combination with NPs enabled good discrimination of TTS from ACS; (5) an ‘optimal’ troponin value of ≥25.92 times the ULN identifies ACS with high clinical probability.

### Troponin and BNP as biomarkers in TTS

Patients presenting with acute cardiac chest pain and ECG changes should undergo rapid assessment for ACS with consideration of urgent diagnostic coronary angiography. However, utility of cardiac biomarkers remains in cases of diagnostic uncertainty, or as a parallel line of investigation. Chest pain represents one of the most frequent presentations to emergency services and therefore ACS represents a common and important differential diagnosis.[44] The 4^th^ Universal Definition of Myocardial Infarction classifies TTS as a form of myocardial injury, with high catecholamine surges known to trigger troponin release from cardiomyocytes, and coronary microvascular dysfunction, high myocardial strain hypercontractility, and high ventricular afterload each being present in TTS which may contribute to an element of myocardial ischaemia.[45] Owing to the high sensitivity and diagnostic accuracy of newer troponin assays,[46] where primary percutaneous coronary intervention is not indicated, rapid ‘rule-in’ and ‘rule-out’ algorithms are used to reduce delays in diagnosis and discharge/treatment for ACS. However, European Society of Cardiology (ESC) guidelines for ACS[47,48] or acute heart failure[49] do not give advice to assist in the diagnosis of TTS (or its differentiation from ACS), and it remains a diagnosis of exclusion. The higher troponin concentrations measured in ACS, when compared to TTS (Figure 2A) reflects the extent of myocyte damage present in each condition, with an even larger difference present on subgroup analysis for STEMI (Figure 3A). This more modest increase in troponin concentration is present in TTS despite grossly, albeit mostly temporary abnormal LV contractile function.

Elevation of serum cardiac NPs (BNP or NT-proBNP) is frequent in the acute phase of TTS, consequent to increased ventricular myocardial strain, regional wall motion abnormality and direct catecholamine-induced NP release.[1] The finding that BNP is significantly higher in TTS than ACS (570 patients versus 575 respectively, Figure 2B) is therefore unsurprising. The lack of change on subgroup analysis by ACS type (Figure 3B) likely reflects the equivocal magnitude of myocardial dysfunction in NSTEMI compared to STEMI. Interestingly, the absolute magnitude of change in NT-pro-BNP was significantly greater than BNP (p=0.01, Supplementary Figure 2A), however this difference was reversed on standardization with the difference in BNP trending to be higher than NT-pro-BNP (P=0.05, Supplementary Figure 2B). NT-pro-BNP has previously been suggested to increase in greater increment with NYHA score in heart failure and have higher power to detect modest reductions in LVEF when compared to BNP.[50] These differences between biomarker isoforms likely result from differences in the kinetics of BNP/NT-pro-BNP generation, release and clearance.

### Selection of optimal biomarker values

The ratio of NP to troponin has previously been proposed to differentiate TTS from STEMI.[8] Doyen et al. present BNP to TnI ratios of 642.0 pg/μg (331.8 to 1226.5 pg/μg) in TTS compared to 184.5 pg/μg (50.5 to 372.3 pg/μg) for NSTEMI and 7.5 pg/μg (2.0 to 29.6 pg/μg) for STEMI (p<0.001 for both).[25] Budnik et al. similarly present a ratio of NT-pro-BNP to TnI ratio of 2235.2 (1086.2 to 9480.8) for TTS *versus* 81.6 (47.9 to 383.3) for STEMI (p<0.001).[21] Finally, Randhawa et al. provide a ratio of BNP to TnT of 1,292.1 (443.4 to 2,657.9) for TTS and 226.9 (69.91-426.32) for AMI (p < 0.001).[36] Unfortunately, it is not possible to create a standardized ratio on this information alone due to the differences in biomarker isoform and assays used, and lacking of patient level data.

Whilst these studies suggest clear benefit to the clinical use of NP to troponin ratio, based on the 13 studies that reported paired values for troponin and NPs, ROC curve analysis demonstrated there was no additional value of combining NP with cTn over cTn alone, as baseline discriminatory power from cTn is higher (Supplementary Figure 3). This also suggested that optimal biomarker thresholds could not be selective based on ROC curves alone. The Youden index is often used to determine an ‘optimal’ point, however, it is not useful in this context as cannot appreciate the different ‘weight’ one might ascribe to prioritising a certain clinical diagnosis, or where differences in prevalence are present. In this case, sensitivity of diagnosing ACS should be valued higher than any ‘optimal’ cut-off, and initiation of therapy for ACS should not be delayed over a possible TTS diagnosis. The Youden index would attribute equal weights to sensitivity and specificity and therefore accept missing a STEMI over misclassifying a TTS.

An alternative approach that incorporates both pre-test likelihood and pre-defined performance goals was utilising recursive partitioning and regression tree analysis.[18] This enables for a ‘prior’ probability distribution to account for the increased prevalence of ACS when compared to TTS. Given early estimates suggested 1–2% of patients with suspected ACS are eventually diagnosed with TTS[3,4] and more recent estimates report TTS constitute up 6% of female patients presenting with suspected STEMI,[5] we used a conservative prior assumption that 90% of cases would ultimately be diagnosed as ACS, to reflect that TTS is still ultimately an overlooked condition.

When the decision tree considered all available data for both NPs and cTn, a threshold value for cTn alone of ≥25.92 times the ULN made ACS far more likely – with a probability exceeding 95%. This provided a systematic and unbiased method of identifying the ‘optimal’ value for the differentiation of the 2 conditions; and demonstrates that troponin alone is the superior biomarker for diagnostic discrimination in this context.

Translating this to ‘real-world’ assay concentrations, this represents values of 492.5ng/L for Abbott Architect hs-TnI assay and 414.7ng/L for Roche Cobas hs-TnT. Overall, this threshold yielded sensitivity and specificity values for troponin of 85.71% and 53.57% respectively, with a PPV of 94.32% and NPV of 29.40%. This seems adequately powered to discern ACS and TTS above these troponin concentrations; below, however, NPV is too low for troponin to provide a meaningful clinical differentiation.

### Novel biomarkers for TTS

This study further highlights the lack of specific biomarkers for TTS and illustrates the need for novel biomarkers. In brief, non-coding RNA (ncRNA) represent novel disease biomarkers as ncRNA expression profiles change specifically in various disease states.[51] microRNAs (miR) −1, miR-16, miR-26a and miR-133a are all significantly raised in TTS *versus* healthy control,[42] and the pattern of change was significantly different to STEMI. miR-1 and miR-133a were higher in STEMI, whereas miR-16 and miR-26a were raised only in TTS.[42] Interestingly, miR-16 and miR-26a have recently been identified to be involved in the pathophysiology of TTS.[52]

Given the pathophysiological involvement of inflammation and microvascular dysfunction in TTS,[53–55] markers of inflammation and vascular stress response have been investigated for use as diagnostic tools in TTS. IL-2, IL-4, IL-10, IFN-γ and TNF-α were observed to be higher in patients with acute TTS *versus* AMI, and IL-6 higher in AMI.[38] The two vasoconstricting peptides co-peptin and endothelin are differentially changed in TTS *versus* AMI. Co-peptin levels are normal or marginally elevated in TTS compared to more substantial elevations in patients with AMI,[22,23,27] whereas endothelin levels are increased to a similar degree in TTS and STEMI.[42] Glycocalyx levels also seem significantly elevated in Takotsubo compared to healthy controls.[56] The mechanisms underlying these changes has recently been reviewed elsewhere.[9] Taken together, this highlights the need for further work within this field to identify more specific biomarkers than enables the rapid triage of patients with ACS and TTS.

### Limitations

There are several limitations present of our study. Firstly, research in TTS is largely retrospective, and therefore the trials included within this analysis may have been subject to sampling bias. Due to the paucity of randomized control trials in TTS, data was standardized to a factor of the ULN and pooled for total NPs and total troponin. Large repositories such as interTAK[2] were used to strengthen the data. However, inclusion of a wide variety of study designs lead to increased study heterogeneity, which is seen throughout the data presented, and was largely unaffected on sensitivity analysis by exclusion of studies based on study design, timing of biomarker or presence of bias. Further, approximately only half of the included studies included the paired comparison of troponin and NPs. This limited the ability to create a standardized NP to troponin ratio that could undergo ROC curve analysis. Finally, studies frequently did not clarify the biomarker assay used and corresponding authors often did not respond to requests for clarification. We substituted averaged assay values from standard populations[13,14] to attempt to compensate for this, however, we would have preferred to use local assay specific thresholds.

## Conclusion

Whilst no substitute for coronary angiography, cardiac biomarkers represent a useful tool in routine clinical diagnosis. Troponin is significantly lower, and NPs are significantly higher in TTS compared to ACS. Troponin had greater power than NPs at discriminating TTS and ACS, and a threshold value of ≥26 times the ULN for troponin was found to identify patients as extremely likely to have ACS.

## Funding

This work was further supported by grants from the Medical Research Council (London, UK) (G1000737), Guy’s and St Thomas’ Charity (London, UK; R060701, R100404), British Heart Foundation (Birmingham, London; TG/15/1/31518, FS/15/13/31320), and the UK Department of Health through the National Institute for Health Research Biomedical Research Centre at Guy’s and St Thomas’ NHS Trust. ARL is supported by the Fondation Leducq Network of Excellence in Cardio-Oncology and the Royal Brompton Cardio-Oncology Centre of Excellence is supported by the The Big Heart Foundation. LSC is funded through an NIHR Academic Clinical Fellowship and TEK is funded through an NIHR clinical lectureship (CL-2019-17-006).

## Conflict of interest

ARL has received speaker, advisory board or consultancy fees and/or research grants from Janssens-Cilag Ltd, Astellas Pharma, Pfizer, Novartis, Servier, Astra Zeneca, Bristol Myers Squibb, GSK, Amgen, Takeda, Roche,, Clinigen Group, Eli Lily, Eisai Ltd, Ferring Pharmaceuticals, Boehringer Ingelheim, Akcea Therapeutics, Myocardial Solutions, iOWNA Health and Heartfelt Technologies Ltd.

## Data availability statement

Available on request.

## References

[1] Lyon AR, Bossone E, Schneider B, Sechtem U, Citro R, Underwood SR, et al. Current state of knowledge on Takotsubo syndrome: a Position Statement from the Taskforce on Takotsubo Syndrome of the Heart Failure Association of the European Society of Cardiology. Eur J Heart Fail 2016;18:8–27. 10.1002/ejhf.424.

[2] Templin C, Ghadri JR, Diekmann J, Napp LC, Bataiosu DR, Jaguszewski M, et al. Clinical Features and Outcomes of Takotsubo (Stress) Cardiomyopathy. New England Journal of Medicine 2015;373:929–38. 10.1056/NEJMoa1406761.

[3] Gianni M, Dentali F, Grandi AM, Sumner G, Hiralal R, Lonn E. Apical ballooning syndrome or takotsubo cardiomyopathy: a systematic review. Eur Heart J 2006;27:1523–9. 10.1093/EURHEARTJ/EHL032.

[4] Kurowski V, Kaiser A, Von Hof K, Killermann DP, Mayer B, Hartmann F, et al. Apical and midventricular transient left ventricular dysfunction syndrome (tako-tsubo cardiomyopathy): frequency, mechanisms, and prognosis. Chest 2007;132:809–16. 10.1378/CHEST.07-0608.

[5] Redfors B, Vedad R, Angerås O, Råmunddal T, Petursson P, Haraldsson I, et al. Mortality in takotsubo syndrome is similar to mortality in myocardial infarction - A report from the SWEDEHEART. Int J Cardiol 2015;185:282–9. 10.1016/j.ijcard.2015.03.162.

[6] Madhavan M, Borlaug BA, Lerman A, Rihal CS, Prasad A. Stress hormone and circulating biomarker profile of apical ballooning syndrome (Takotsubo cardiomyopathy): Insights into the clinical significance of B-type natriuretic peptide and troponin levels. Heart 2009;95:1436–41. 10.1136/hrt.2009.170399.

[7] Stiermaier T, Santoro F, Graf T, Guastafierro F, Tarantino N. Prognostic value of N-Terminal Pro-B-Type Natriuretic Peptide in Takotsubo syndrome. Clinical Research in Cardiology 2018;0:0. 10.1007/s00392-018-1227-1.

[8] Fröhlich GM, Schoch B, Schmid F, Keller P, Sudano I, Lüscher TF, et al. Takotsubo cardiomyopathy has a unique cardiac biomarker profile: NT-proBNP/myoglobin and NT-proBNP/troponin T ratios for the differential diagnosis of acute coronary syndromes and stress induced cardiomyopathy. Int J Cardiol 2012;154:328–32. 10.1016/j.ijcard.2011.09.077.

[9] Khan H, Gamble D, Mezincescu A, Abbas H, Rudd A, Dawson D. A systematic review of biomarkers in Takotsubo syndrome: A focus on better understanding the pathophysiology. IJC Heart and Vasculature 2021;34:100795. 10.1016/j.ijcha.2021.100795.

[10] Jaguszewski M, Osipova J, Ghadri JR, Napp LC, Widera C, Franke J, et al. A signature of circulating microRNAs differentiates takotsubo cardiomyopathy from acute myocardial infarction. Eur Heart J 2014;35:999–1006. 10.1093/eurheartj/eht392.

[11] Couch LS, Fiedler J, Chick G, Clayton R, Dries E, Wienecke LM, et al. Circulating microRNAs predispose to takotsubo syndrome following high-dose adrenaline exposure. Cardiovasc Res 2022;118:1758–70. 10.1093/cvr/cvab210.

[12] Madhavan M, Prasad A. Proposed Mayo Clinic criteria for the diagnosis of Tako-Tsubo cardiomyopathy and long-term prognosis. Herz 2010;35:240–3. 10.1007/s00059-010-3339-x.

[13] Apple FS, Wu AHB, Sandoval Y, Sexter A, Love SA, Myers G, et al. https://academic.oup.com/cardiovascres/article/118/8/1932/6169160. Clin Chem 2020;66:434–44. 10.1093/CLINCHEM/HVZ029.

[14] BNP, NT-proBNP, and MR-proANP Assays: Analytical Characteristics Designated by Manufacturer IFCC Committee on Clinical Applications of Cardiac Bio-Markers (C-CB) v082318. International Federation of Clinical Chemistry and Laboratory Medicine 2018:1–5. https://www.ifcc.org/media/477439/bnp-nt-probnp-and-mr-proanp-assays-analytical-characteristics-designated-by-manufacturer-v08232018.pdf%09%09%09%0A (accessed June 6, 2022).

[15] Wan X, Wang W, Liu J, Tong T. Estimating the sample mean and standard deviation from the sample size, median, range and/or interquartile range. BMC Med Res Methodol 2014;14:1–13. 10.1186/1471-2288-14-135/TABLES/3.

[16] The Cochrane Collaboration. Review Manager (RevMan) 2020.

[17] Study Quality Assessment Tools | NHLBI, NIH n.d.:Quality Assessment of Case-Control Studies. https://www.nhlbi.nih.gov/health-topics/study-quality-assessment-tools (accessed October 25, 2023).

[18] Breiman L, Friedman J, Olshen R, Stone C. Classification And Regression Trees. 1st Ed. Wadsworth; 1984. 10.1201/9781315139470.

[19] Ahmed KA, Madhavan M, Prasad A. Brain natriuretic peptide in apical ballooning syndrome (Takotsubo/stress cardiomyopathy): Comparison with acute myocardial infarction. Coron Artery Dis 2012;23:259–64. 10.1097/MCA.0b013e3283526a57.

[20] Ahmed M, Sardana M, Rasla S, Escobar J, Bote J, Iskandar A, et al. Comparative left ventricular mechanical deformation in acute apical variant stress cardiomyopathy and acute anterior myocardial infarction utilizing 2-dimensional longitudinal strain imaging. Echocardiography 2020;37:832–40. 10.1111/echo.14675.

[21] Budnik M, Kochanowski J, Piatkowski R, Wojtera K, Peller M, Gaska M, et al. Simple markers can distinguish Takotsubo cardiomyopathy from ST segment elevation myocardial infarction. Int J Cardiol 2016;219:417–20. 10.1016/j.ijcard.2016.06.015.

[22] Budnik M, Białek S, Peller M, Kiszkurno A, Kochanowski J, Kucharz J, et al. Serum copeptin and copeptin/NT-proBNP ratio - new tools to differentiate takotsubo syndrome from acute myocardial infarction. Folia Med Cracov 2020;60:5–14. 10.24425/FMC.2020.133481.

[23] Burgdorf C, Schubert A, Schunkert H, Kurowski V, Radke PW. Release patterns of copeptin and troponin in Tako-Tsubo cardiomyopathy. Peptides (NY) 2012;34:389–94. 10.1016/J.PEPTIDES.2012.01.022.

[24] Demir SÇ, Demir E, Çatalkaya S. Electrocardiographic and seasonal patterns allow accurate differentiation of Tako-Tsubo cardiomyopathy from acute anterior myocardial infarction: Results of a multicenter study and systematic overview of available studies. Biomolecules 2019;9:51. 10.3390/biom9020051.

[25] Doyen D, Moceri P, Chiche O, Schouver E, Cerboni P, Chaussade C, et al. Cardiac biomarkers in Takotsubo cardiomyopathy. Int J Cardiol 2014;174:798–801. 10.1016/J.IJCARD.2014.04.120.

[26] Hong JY, Ryu SK, Park JY, Park SH, Choi J. Early differentiation of stress cardiomyopathy from acute anterior wall myocardial using changing cardiac enzyme patterns. J Cardiovasc Imaging 2021;29:228–33. 10.4250/jcvi.2020.0167.

[27] Højagergaard MA, Hassager C, Christensen TE, Bang LE, Gøtze JP, Ostrowski SR, et al. Biomarkers in patients with Takotsubo cardiomyopathy compared to patients with acute anterior ST-elevation myocardial infarction. 10.1080/1354750X20191710767 2020;25:137–43. 10.1080/1354750X.2019.1710767.

[28] Möller C, Stiermaier T, Meusel M, Jung C, Graf T, Eitel I. Microcirculation in patients with takotsubo syndrome—the prospective circus-tts study. J Clin Med 2021;10:2127. 10.3390/jcm10102127.

[29] Nascimento FO, Yang S, Larrauri-Reyes M, Pineda AM, Cornielle V, Santana O, et al. Usefulness of the troponin-ejection fraction product to differentiate stress cardiomyopathy from ST-segment elevation myocardial infarction. American Journal of Cardiology 2014;113:429–33. 10.1016/j.amjcard.2013.10.013.

[30] Novo G, Giambanco S, Bonomo V, Sutera MR, Giambanco F, Rotolo A, et al. Troponin I/ejection fraction ratio: A new index to differentiate Takotsubo cardiomyopathy from myocardial infarction. Int J Cardiol 2015;180:255–7. 10.1016/j.ijcard.2014.11.186.

[31] Núñez-Gil IJ, Fernández-Ortiz A, Pérez-Isla L, Luaces M, García-Rubira JC, Vivas D, et al. Clinical and prognostic comparison between left ventricular transient dyskinesia and a first non-ST-segment elevation acute coronary syndrome. Coron Artery Dis 2008;19:449–53. 10.1097/MCA.0b013e32830eab74.

[32] Parkkonen O, Nieminen MT, Vesterinen P, Tervahartiala T, Perola M, Salomaa V, et al. Low MMP-8/TIMP-1 reflects left ventricle impairment in takotsubo cardiomyopathy and high TIMP-1 may help to differentiate it from acute coronary syndrome. PLoS One 2017;12:e0173371. 10.1371/journal.pone.0173371.

[33] Patel SM, Lennon RJ, Prasad A. Regional wall motion abnormality in apical ballooning syndrome (Takotsubo/stress cardiomyopathy): Importance of biplane left ventriculography for differentiating from spontaneously aborted anterior myocardial infarction. International Journal of Cardiovascular Imaging 2012;28:687–94. 10.1007/s10554-011-9911-5.

[34] Pawlak M, Roik M, Kochanowski J, Scisło P, Kowalik R, Huczek Z, et al. Comparison of on-admission ST-segment elevation tako-tsubo patients and myocardial infarction women: in-hospital course and long-term follow-up - PubMed. Kardiol Pol 2012;70:233–40.

[35] Pirlet C, Pierard L, Legrand V, Gach O. Ratio of high-sensitivity troponin to creatine kinase-MB in takotsubo syndrome. Int J Cardiol 2017;243:300–5. 10.1016/j.ijcard.2017.05.107.

[36] Randhawa MS, Dhillon AS, Taylor HC, Sun Z, Desai MY. Diagnostic utility of cardiac biomarkers in discriminating takotsubo cardiomyopathy from acute myocardial infarction. J Card Fail 2014;20:2–8. 10.1016/j.cardfail.2013.12.004.

[37] Samul-Jastrzębska J, Roik M, Wretowski D, Łabyk A, Ślubowska A, Bizoń A, et al. Evaluation of the intertak diagnostic score in differentiating takotsubo syndrome from acute coronary syndrome. A single center experience. Cardiol J 2021;28:416–22. 10.5603/CJ.a2019.0086.

[38] Santoro F, Costantino MD, Guastafierro F, Triggiani G, Ferraretti A, Tarantino N, et al. Inflammatory patterns in Takotsubo cardiomyopathy and acute coronary syndrome: A propensity score matched analysis. Atherosclerosis 2018;274:157–61. 10.1016/j.atherosclerosis.2018.05.017.

[39] Sharkey SW, Lesser JR, Menon M, Parpart M, Maron MS, Maron BJ. Spectrum and Significance of Electrocardiographic Patterns, Troponin Levels, and Thrombolysis in Myocardial Infarction Frame Count in Patients With Stress (Tako-tsubo) Cardiomyopathy and Comparison to Those in Patients With ST-Elevation Anterior Wall Myoc. American Journal of Cardiology 2008;101:1723–8. 10.1016/j.amjcard.2008.02.062.

[40] Tarantino N, Santoro F, Di Biase L, Di Terlizzi V, Vitale E, Barone R, et al. Chromogranin-A serum levels in patients with takotsubo syndrome and ST elevation acute myocardial infarction. Int J Cardiol 2020;320:12–7. 10.1016/j.ijcard.2020.07.040.

[41] Zorzi A, Baritussio A, ElMaghawry M, Siciliano M, Migliore F, Perazzolo Marra M, et al. Differential diagnosis at admission between Takotsubo cardiomyopathy and acute apical-anterior myocardial infarction in postmenopausal women. Eur Heart J Acute Cardiovasc Care 2016;5:298–307. 10.1177/2048872615585515.

[42] Jaguszewski M, Osipova J, Ghadri JR, Napp LC, Widera C, Franke J, et al. A signature of circulating microRNAs differentiates takotsubo cardiomyopathy from acute myocardial infarction. Eur Heart J 2014;35:999–1006. 10.1093/eurheartj/eht392.

[43] Topf A, Mirna M, Paar V, Motloch LJ, Grueninger J, Dienhart C, et al. The differential diagnostic value of selected cardiovascular biomarkers in Takotsubo syndrome. Clinical Research in Cardiology 2022;111:197–206. 10.1007/s00392-021-01956-2.

[44] NHS England. Hospital Accident & Emergency Activity 2020-21 - NHS Digital. NHS Digital 2021:data and information. https://digital.nhs.uk/data-and-information/publications/statistical/hospital-accident--emergency-activity/2020-21 (accessed October 4, 2021).

[45] Thygesen K, Alpert JS, Jaffe AS, Chaitman BR, Bax JJ, Morrow DA, et al. Fourth Universal Definition of Myocardial Infarction (2018). J Am Coll Cardiol 2018;72:2231–64. 10.1016/j.jacc.2018.08.1038.

[46] Anand A, Shah ASV, Beshiri A, Jaffe AS, Mills NL. Global adoption of high-sensitivity cardiac troponins and the universal definition of myocardial infarction. Clin Chem 2019;65:484–9. 10.1373/clinchem.2018.298059.

[47] Collet J-P, Thiele H, Barbato E, Barthélémy O, Bauersachs J, Bhatt DL, et al. 2020 ESC Guidelines for the management of acute coronary syndromes in patients presenting without persistent ST-segment elevation. Eur Heart J 2021;42:1289–367. 10.1093/eurheartj/ehaa575. Erratum in: Eur Heart J 2021;42:2298. doi: 10.1093/eurheartj/ehab285.

[48] Ibanez B, James S, Agewall S, Antunes MJ, Bucciarelli-Ducci C, Bueno H, et al. 2017 ESC Guidelines for the management of acute myocardial infarction in patients presenting with ST-segment elevation. Eur Heart J 2018;39:119–77. 10.1093/eurheartj/ehx393.

[49] McDonagh TA, Metra M, Adamo M, Gardner RS, Baumbach A, Böhm M, et al. 2021 ESC Guidelines for the diagnosis and treatment of acute and chronic heart failure. Eur Heart J 2021;42:3599–726. 10.1093/eurheartj/ehab368.

[50] Seino Y, Ogawa A, Yamashita T, Fukushima M, Ogata KI, Fukumoto H, et al. Application of NT-proBNP and BNP measurements in cardiac care: A more discerning marker for the detection and evaluation of heart failure. Eur J Heart Fail 2004;6:295–300. 10.1016/j.ejheart.2003.12.009.

[51] de Gonzalo-Calvo D, Vea A, Bär C, Fiedler J, Couch LS, Brotons C, et al. Circulating non-coding RNAs in biomarker-guided cardiovascular therapy: a novel tool for personalized medicine? Eur Heart J 2019;40:1643–50. 10.1093/EURHEARTJ/EHY234.

[52] Couch LS, Fiedler J, Chick G, Clayton R, Dries E, Wienecke LM, et al. Circulating microRNAs predispose to takotsubo syndrome following high-dose adrenaline exposure. Cardiovasc Res 2022;118:1758–70. 10.1093/cvr/cvab210.

[53] Omerovic E, Citro R, Bossone E, Redfors B, Backs J, Bruns B, et al. Pathophysiology of Takotsubo Syndrome – a joint scientific statement from the Heart Failure Association Takotsubo Syndrome Study Group and Myocardial Function Working Group of the European Society of Cardiology – Part 1: Overview and the central role for. Eur J Heart Fail 2021:Placeholder reference.

[54] Omerovic E, Citro R, Bossone E, Redfors B, Backs J, Bruns B, et al. Pathophysiology of Takotsubo syndrome – a joint scientific statement from the HFA TTS and Myocardial Function Working Group of the ESC – Part 2: vascular pathophysiology, gender and sex hormones, genetics, chronic cardiovascular problems and clinical impl. Eur J Heart Fail 2022;24:274–86. 10.1002/ejhf.2368.

[55] Couch LS, Channon K, Thum T. Molecular Mechanisms of Takotsubo Syndrome. Int J Mol Sci 2022;23:12262. 10.3390/IJMS232012262.

[56] Nguyen TH, Liu S, Ong GJ, Stafford I, Frenneaux MP, Horowitz JD. Glycocalyx shedding is markedly increased during the acute phase of Takotsubo cardiomyopathy. Int J Cardiol 2017;243:296–9. 10.1016/j.ijcard.2017.04.085.

